# Transmissibility of 2019 Novel Coronavirus: zoonotic vs. human to human transmission, China, 2019-2020

**DOI:** 10.1101/2020.03.16.20037036

**Authors:** Kenji Mizumoto, Katsushi Kagaya, Gerardo Chowell

**Affiliations:** Graduate School of Advanced Integrated Studies in Human Survivability, Kyoto University Yoshida-Nakaadachi-cho, Sakyo-ku, Kyoto, Japan; Hakubi Center for Advanced Research, Kyoto University, Yoshidahonmachi, Sakyo-ku,Kyoto, Japan; Department of Population Health Sciences, School of Public Health, Georgia State University, Atlanta, Georgia, USA; Seto Marine Biological Laboratory, Field Science, Education and Reseach Center, Kyoto University, Shirahama-cho, Nishimuro-gun, Wakayama 649-2211 Japan

## Abstract

**Objectives:** The novel coronavirus (2019-nCoV) originating from Wuhan has rapidly spread throughout China. While the origin of the outbreak remains uncertain, accumulating evidence links a wet market in Wuhan for the early spread of 2019-nCoV. Similarly, the influence of the marketplace on the early transmission dynamics is yet to be investigated.

**Methods:** Using the daily series of 2019-nCov incidenceincluding contact history with the market, we have conducted quantitative modeling analyses to estimate the reproduction numbers (*R*) for the market-to-human and human-to-human transmission together with the reporting probability and the early effects of public health interventions.

**Results:** Our mean *R* estimates for China in 2019-2020 are estimated at 0.37 (95%CrI: 0.02-1.78) for market-to-human transmission, and 3.87 (95%CrI: 3.18-4.78) for human-to-human transmission, respectively. Moreover we estimated that the reporting rate cases stemming from market-to-human transmission was 3-31 fold higher than that for cases stemming from human-to-human transmission, suggesting that contact history with the wet market played a key role in identifying 2019-nCov cases.

**Conclusions:** Our findings suggest that the proportions of asymptomatic and subclinical patients constitute a substantial component of the epidemic’s magnitude. Findings suggest that the development of rapid diagnostic tests could help bring the epidemic more rapidly under control.

## Introduction

A novel coronavirus (SARS-CoV-2) originating from Wuhan has rapidly spread throughout China, with multiple cases exported to 25 countries around the world. The cumulative number of confirmed cases has reached 75204 including 2006 deaths as of February 19, 2020 [1]. Early mean estimates of the reproduction number have been estimated in the range 1.4-2.5, comparable with estimates for seasonal, 2009 pandemic flu, SARS and MERS [2-5]. The epidemic continues to spread throughout China at an alarming rate while chains of transmission start to unfold in other countries including Singapore, Korea, and Japan, indicating that the possibility of a pandemic scenario cannot be ruled out.

Evidence suggest that the novel coronavirus likely jumped from a primary reservoir (e.g, horseshoe bats) to an intermediary reservoir, possibly generating an outbreak among wild animals in at least one wet market in Wuhan, China [6-7]. The virus first infected multiple individuals visiting the Huanan Seafood Wholesale Market, initiating multiple chains of transmission that ensured sustained transmission in the human population [8]. While the detailed origin of the outbreak remains uncertain, significant evidence strongly links the Huanan Seafood Wholesale Market in Wuhan for the early spread of the novel coronavirus (COVID-19) among humans [7].

In this paper we conduct quantitative modeling analyses to quantify the role of the wet marketplace on the early transmission dynamics of the novel coronavirus in China [9]. For this purpose, we analyzed and modeled data that stratifies the market hazard (market-to-human transmission) and human-to-human transmission.

## Materials and methods

### Epidemiological incidence cases

Daily series of laboratory-confirmed COVID-19 cases were extracted from a recently published study [7]. From December 8, 2019 to January 21, 2020, we analyzed a total of 425 confirmed cases by date of symptoms onset including information on whether the case was linked to the Huanan Seafood Wholesale Market. That is, this unique case series stratifies cases with visiting history to the Huanan Seafood Wholesale Market, which has been purported as source of this large epidemic [9], and those arising from human-to-human transmission. Because the corresponding incidence curve is subject reporting delays, the last 12 epidemic days were excluded from our analysis. Thus, the study period is set from December 8^th^, 2019 to January 9^th^, 2020.

### Epidemiological modelling

We model two types of infections: a) market hazard (market-to-human transmission, including primary infections arising from zoonotic transmission to some extent) and b) human-to-human transmission chains. A model schematic of the transmission dynamics is provided in Figure 1. Primary infection (index case) occurs as a result of a spillover event, and primary infections may generate secondary cases. The number of infected individuals stemming from the market-to-human and the human-to-human route are denoted by *i*_*m*_ and *i*_h_, respectively.

**Figure 1.**
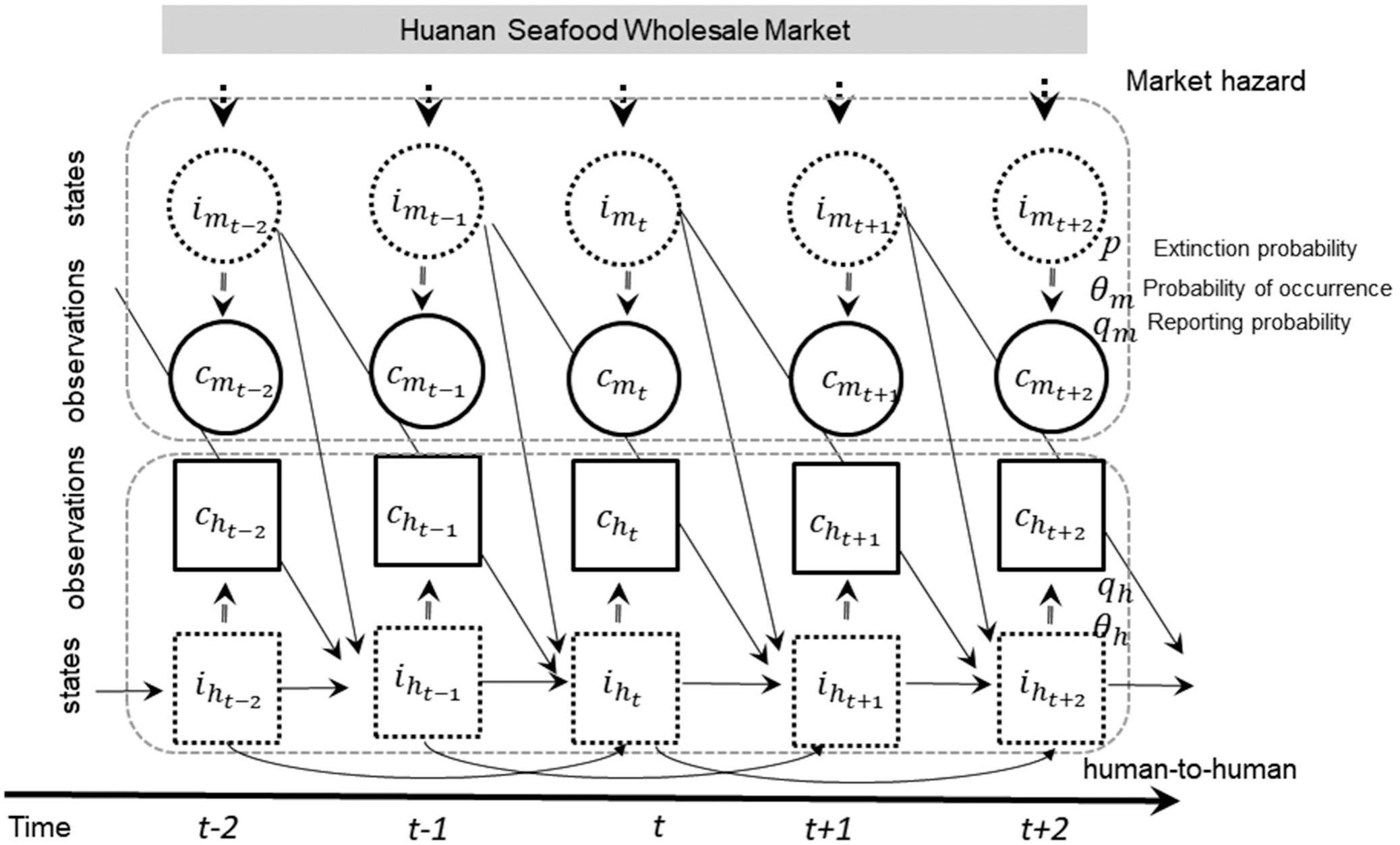
Schematic model diagram of COVID-19 transmission dynamics. A schematic model diagram of transmission dynamics is presented. The transmission routes are classified into market-to-human transmission and human-to-human transmission. Infected through market-to-human route and human-to-human route is indicated by *i*_*m*_ and *i*_h_, respectively. Reported cases of primary case infected through market-to-human route and of secondary or later cases though human-to-human route is indicated by *c*_*m*_ and *c*_h_, respectively. A virus with substantial human-to-human transmission potential could yield subsequent transmissions caused by human-to-human transmission.

We employed a discrete-time integral equation to capture the daily incidence series with contact history. Let *f*_s_ denote the probability mass function of the serial interval of length *s* days, which is given by

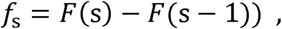

For *s* >0 where *F*(.) represents the cumulative distribution function of the gamma distribution. The expected number of new cases with onset day *t* after secondary infections, E[*c*_*h*_(t)], is written as follows,

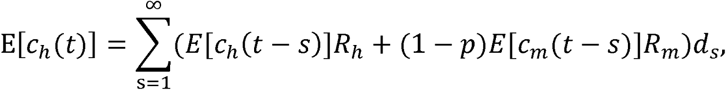

where E[*c*_*m*_(t)] represents the expected number of new cases with onset day *t* infected through market-to-human transmission, *p* is the conditional probability of extinction within one generation, given by 1/(*R*_*m*_+1) [10-11], where *R*_h_ and *R*_*m*_ represents the average number of secondary infections generated by one single infection generated from the market-to-human or from the human-to-human transmission, respectively.

To account for the type of infection dependent probability of occurrence, *θ* _*j*_ [12], we assume that the number of observed cases for infection type *j* on day *t, h*_j_(*t*), occurred according to a Bernoulli sampling process, with the expected values E(*c*_j_;*H*_t-1_), where E(*c*_j_; *H*_t-1_) denotes the conditional expected incidence, on day *t*, given the history of observed data from day 0 to day (*t*-1), denoted by *H*_t-1_. Thus, the number of expected newly observed cases is written as follows:

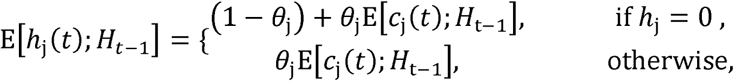

Subsequently, we also account for the reporting probability that depends on the type of infection. We assume that the number of reported cases by infection type *j* on day *t, h*_j_(*t*), is the product of type-dependent reporting rate, *q*_j_, and the actual number of cases, *c*_j_(t).

Further, we model the time-dependent variation in the transmissibility associated with implementation of public interventions in Wuhan. This time dependence was modelled by introducing a parameter *δ*_ti_, by transmission route. Time dependence in primary infection and secondary infection is given by

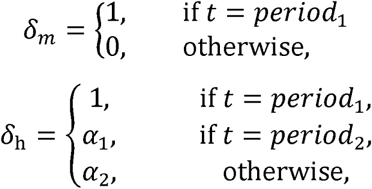

where *α*_*1*_ *and α*_*2*_ scale the intensity of public health interventions (where *α*_*1*_ and *α*_*2*_ is expected to be smaller than 1) and period_1_ and period_2_ define the study periods according to timing of the closing of the Huanan Seafood Wholesale Market (December 31 in 2019) and the date on which the novel coronavirus was officially declared as the causative pathogen of the outbreak by China CDC (January 8 in 2020) [8]. After the Huanan Seafood Wholesale Market was closed on December 31 in 2019, we assume the market hazard is negligible

The number of expected newly observed cases should be updated as

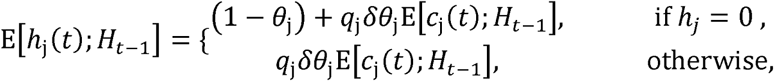

We assume the incidence, *h*_j_ is the result of the Poisson sampling process with the expected value of E[*h*_j_]. The likelihood function for the time series of observed cases of primary infection through the market-to-human transmission and secondary infection through human-to-human transmission that we employ to estimate the reproduction number and other relevant parameters is given by:

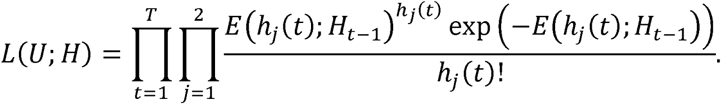

where *U* indicates parameter sets that are estimated from this likelihood.

The serial interval is assumed to follow a gamma distribution with the mean and SD at 7.5 and 3.4 days, respectively, based on ref. [8]. The maximum value of the serial interval was fixed at 16 days as the cumulative probability distribution of the gamma distribution up to 16 days reaches 0.97.

For sensitivity analyses, we examined the effect of varying the mean of the serial interval on *R*_j_ by varying the mean serial interval from 5.5 to 9.5 days.

We estimated model parameters and made projections using a Monte Carlo Markov Chain (MCMC) method in a Bayesian framework. Point estimates and the corresponding 95% credibility intervals were drawn from the posterior probability distributions. All statistical analyses were conducted in R version 3.5.2 (R Foundation for Statistical Computing, Vienna, Austria) and the ‘rstan’ package (No-U-Turn-Sampler (NUTS)).

## Results

The daily series of nCov incidence in China in 2019-2020 are displayed in Figure 2. Visual inspection indicates that the trend of nCov laboratory-confirmed cases that are not linked to the Huanan Seafood Wholesale Market follows a rapid growth pattern, whereas those cases linked to the Huanan Seafood Wholesale Market follow an overall stable incidence pattern until early January in 2020, likely associated with the closing of the Huanan Seafood Wholesale Market in Wuhan.

**Figure 2.**
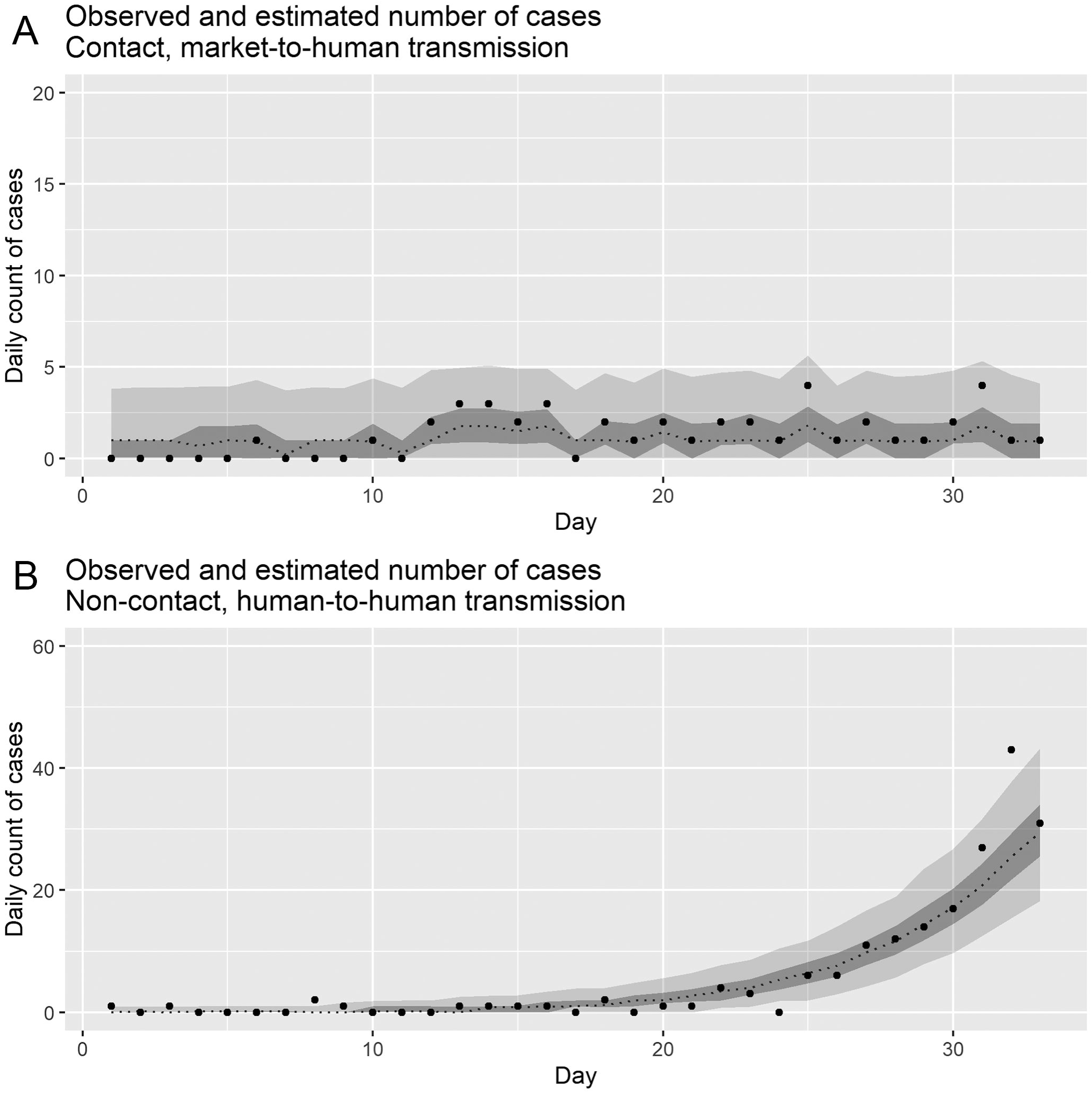
Observed and posterior estimates of the daily number of the COVID-19 cases in China, 2019-2020. Total number of cases with or without contact history with the Huanan Seafood Wholesale Market for our study period is 43 and 187, respectively. Observed laboratory-confirmed cases are the dots. Dashed lines indicates 50 percentile, and areas surrounded by light grey and deep grey indicates 95% and 50% credible intervals (CrI) for posterior estimates, respectively. Upper panel (A) correspond to incidence arising from market to human transmission. Lower panel (B) correspond to observed cases infected through human-to-human transmission, respectively. Epidemic day 1 corresponds to the day that starts at December 8, 2020.

Figure S1 displays the observed and model-based posterior estimates of the daily number of nCov laboratory-confirmed cases by transmission route in China from December, 2019 to January, 2020. Overall, our dynamic models are able to capture the temporal dynamics (i.e. incidence) for both market-to-human and human-to-human transmission routes. Importantly, our posterior estimates of the basic reproduction numbers for China in 2019-2020 are estimated at 0.37 (95%CrI: 0.02-1.78) for market-to-human, and 3.87 (95%CrI:3.18-4.78) for human-to-human, respectively. Other parameter estimates for the probability of occurrence and reporting rate are estimated at 0.93 (95% CrI: 0.74-1.00) and 0.028 (95% CrI: 0.02-0.04) for market-to-human and 0.90 (95% CrI: 0.73-0.99) and 0.00 (95% CrI: 0.00-0.01) for human-to-human, respectively. A Tukey-Kramer test indicated significant differences in probabilities of occurrence (p <0.001) and reporting probability (p <0.001) according to infection route. Moreover, the proportionate reduction in transmissibility brought about the enhanced public health interventions is estimated to be 0.07 (95% CrI: 0.00-0.28) after January 1, and to be 0.50 (95% CrI: 0.02-0.97) after January 8, 2020, which is measured using a time-dependent scaling factor, *α*.

The total number of estimated laboratory-confirmed cases (i.e. cumulative incidence) are 43.0 (95% CrI: 26.4-63.7) for cases infected though market-to-human transmission, 174.8 (95% CrI: 134.3-216.7) for cases infected through human-to-human transmission. For comparison, the actual numbers of reported laboratory-confirmed cases during our study period are 43 and 187, respectively.

Furthermore, we inferred the total number of nCov infections by transmission route to quantify the nCov burden in China during our study period. Our results indicate that the total number of unobserved cases (i.e. cumulative infections) by subgroup are 1685.1 (95% CrI: 1375.3- 2009.2) for those cases infected though market-to-human transmission, and 59986.9 (95% CrI: 19925.4-200988.9) for those cases infected through human-to-human transmission.

Results of sensitivity analyses on the mean serial interval ranging from 5.5 to 9.5 are shown in Figures 3. We found that estimates of *R* from human-to-human transmission are somewhat sensitive to changes in the mean serial interval, ranging from 2.63 (95% CrI: 2.31-3.01) to 5.69 (95% CrI: 4.43-7.44), while *R* for primary infections takes a relatively similar range below the epidemic threshold of 1.0.

**Figure 3.**
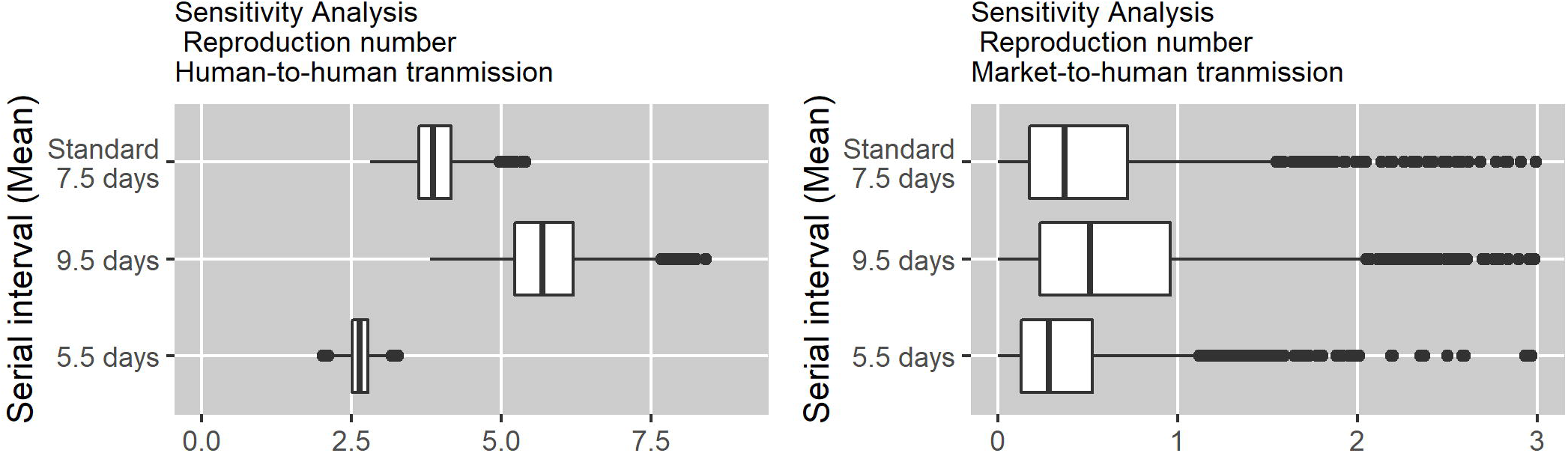
Sensitivity analysis of serial interval on reproduction number by infection route.

## Discussion

We have conducted statistical modeling analyses to elucidate the early transmission characteristics of the novel COVID-19 outbreak in China from December in 2019 to January 2020 by stratifying cases according to transmission route. In particular, we report estimates of the reproduction numbers for market-to-human and human-to-human transmission as well as the occurrence reporting probability and estimate the early effects of public health interventions.

Our posterior estimates of basic reproduction numbers for China in 2019-2020 are estimated to be 0.37 (95%CrI: 0.02-1.78) for market-to-human transmission, and 3.87 (95%CrI: 3.18-4.78) for human-to-human transmission, respectively. These *R* estimates that account for more details of the underlying transmission dynamics shed light on the impact of the COVID-19 epidemic. Thus, our analysis suggests that the total number of unobserved cases (i.e. cumulative infections) are estimated to be 1685.1 (95% CrI: 1375.3- 2009.2) for unobserved cases from market-to-human transmission and 59986.9 (95% CrI: 19925.4-200988.9) for unobserved cases from human-to-human transmission. Our *R* estimates accounting for the unobserved cases (infections) makes the estimated values somewhat higher than previous estimates based solely on observed cases [2, 7]. More importantly, our results indicate a substantial contribution of mild and asymptomatic cases to the epidemic, and this could influence estimates of *R* derived from a single epidemic curve of observed reported cases, which does not fully capture the transmission dynamics [13,14].

Moreover, we also identified a remarkable decrease in the reproduction number, and this decline is in line with the timing of the closing of the Huanan Seafood Wholesale Market (identified as the origin of the epidemic) and the official announcement about the epidemic by China CDC. We found that *R* for human-to-human transmission decreased from 3.87 (95%CrI: 3.18-4.78) to 3.53 (95%CrI: 2.84-4.35) (Relative risk reduction (RRR): 0.07 (95%CrI: 0.00-0.28)) and to 1.91 (95%CrI: 0.10-3.92) (RRR: 0.50 (95%CrI: 0.02-0.97)). However, these estimates need to be interpreted with epidemic context in mind. Specifically, the first intervention targeting market-to-human transmission is unlikely to have mitigated the risk of secondary infections arising from the unfolding transmission chains. The latter decrease is likely the result of enhanced public interventions. However, in the context of rapid and widespread national transmission, efforts to mitigate further spread were complicated by difficulties to track multiple chains of transmission involving cases of the disease with a wide spectrum of illness including asymptomatic and subclinical cases.

Furthermore, reporting probabilities for cases stemming from market-to-human and human-to-human cases were estimated to be low, but we estimated that the reporting rate for market-to-human route was 3-31 fold higher, suggesting that contact history with the Huanan Seafood Wholesale Market played a key role in identifying cases with COVID-19. Indeed, during the initial disease stages, infected individuals with COVID-19 exhibit clinical features that are similar to those of other common respiratory diseases [15]. Moreover, in the context of the ongoing influenza season, the absence of rapid diagnostic testing for this novel disease complicate case identification for this novel coronavirus [16]. Because polymerase chain reaction (PCR) tests, which are time consuming, are not widely available, there is a crucial need to develop rapid diagnostic tests in order to improve the accuracy and speed of clinical diagnosis and enhance reporting rates.

Our results are not free from limitations. First, the serial interval for COVID-19 assumed in our analyses was extrapolated from a previous study, and had to exclude recent data from the epidemic curve due to reporting delays. In fact, our sensitivity analysis on the serial interval indicate some influence on our *R* estimates. Further our *R* estimates could be further improved using additional data collected by field investigations. Second, we cannot rule out the possibility that some cases with contact history with the Huanan Seafood Wholesale Market got infected by contact with humans. However, our low *R* estimate for market-to-human transmission indicates that such a proportion is quite low. Third, our method is a powerful tool to estimate the underlying cases including asymptomatic and mild symptoms, and our results suggests that those proportions constitute a large fraction of the epidemic’s magnitude. Indeed, the result of careful screening and detailed examination of a total of 566 Japanese returnees evacuated from Wuhan city by government-chartered planes during January 29-3, 2020 revealed a ratio of 5 asymptomatic to 4 symptomatic cases [17]. In addition, one study reported that the majority of seasonal coronavirus infections are asymptomatic by most symptom definitions and only 4% of individuals experiencing a seasonal coronavirus infection episode sought medical care for their symptoms [18].

This accumulating evidence further underscores the need to account for a significant fraction of asymptomatic cases. Future serological surveys among the local residents in the epicenter of the epidemic and other areas will shed light into the true magnitude of this epidemic that is having a global impact.

In summary, we have provided epidemiological and transmission parameters of COVID-19 in China from 2019 to 2020 using an ecological model. The power of our approach lies in the ability to infer epidemiological parameters with quantified uncertainty from limited data collected by surveillance systems.

## Data Availability

The datasets generated during and/or analysed during the current study are available from the corresponding author on reasonable request.

## Acknowledgments

KM acknowledges support from the Japan Society for the Promotion of Science (JSPS) KAKENHI Grant Number 18K17368 and from the Leading Initiative for Excellent Young Researchers from the Ministry of Education, Culture, Sport, Science & Technology of Japan. KK acknowledges support from the JSPS KAKENHI Grant Number 18K19336 and 19H05330. GC acknowledges support from NSF grant 1414374 as part of the joint NSF-NIH-USDA Ecology and Evolution of Infectious Diseases program.

## Additional files

**Additional file 1: Figure S1. Posterior estimates of the infections of the COVID-19 in China, 2019-2020**.

Dashed line indicates 50 percentile, and areas surrounded by light grey and deep grey indicates 95% and 50% credible intervals (CrI) for posterior estimates, respectively. Upper panel (A) correspond to infections arising from market-tohuman transmission. Lower panel (B) correspond to infections through human-to-human transmission, respectively. Epidemic day 1 corresponds to the day that starts at December 8,2020.

